# The Association Between Thyroid Dysfunction, Autoimmune Thyroid Disease, and Rheumatoid Arthritis Disease Severity

**DOI:** 10.1101/2022.09.16.22280025

**Authors:** Mohammad Amin Yazdanifar, Mahsa Bagherzadeh-Fard, Mostafa Vahedian, Mohammad Amin Habibi, Mohammad Bagherzadeh, Maryam Masoumi

**Affiliations:** Qom University of Medical Sciences, Qom, Iran

**Keywords:** Rheumatoid arthritis, thyroid dysfunction, anti-thyroid autoantibody, autoimmune thyroid disease, disease activity, disease severity

## Abstract

**Introduction:** Rheumatoid Arthritis (RA) and autoimmune thyroid disease (AITD) are the two most prevalent autoimmune diseases that can coexist due to their similar pathogenesis. Considering the potential effect of AITD on the severity of RA disease, this study aimed to determine the association between thyroid dysfunction, anti-thyroid peroxidase (anti-TPO) positivity, AITD, and RA disease severity in the Iranian population.

**Materials and methods:** Three hundred and fifty RA patients who presented to Shahid Beheshti tertiary care center, Qom, Iran, were included in this cross-sectional study. The data were collected through the patients’ medical records, interviews, physical examinations, and laboratory investigations. The disease activity score in 28 joints for RA with ESR (DAS-28-ESR) was used to divide patients into three groups, remission (DAS-28-ESR ⍰2.6), mild-to-moderate (2.6 < DAS-28-ESR ⍰5.1), and severe disease activity (DAS-28-ESR > 5.1).

**Results:** Using the method explained beforehand, 111, 96, 138 patients were sorted into remission, mild-to-moderate, and severe disease activity groups, respectively. Anti-TPO antibody positivity was 2.93 times more prevalent among patients with higher severity of disease than in remission (OR: 2.93; P-value < 0.001). Patients suffering from a more severe form of the disease were almost 2.7 times more probable to have AITD (OR = 2.71; P-value < 0.001) and 82% more likely to have thyroid dysfunction than the patients in remission (OR = 1.82; P-value = 0.006).

**Conclusions:** It was demonstrated that thyroid dysfunction, anti-TPO antibody positivity, and AITD were significantly more common among RA patients with higher disease activity.

## Introduction

Rheumatoid arthritis (RA) is a chronic systemic autoimmune disease that damages small symmetrical joints. Pathogenesis of the disease is complex and consists of environmental stimulations, such as smoking and dust inhalation, and the immune system malfunction in genetically susceptible individuals (1). Although the hallmark of RA is the articular involvement and the corresponding symptoms, it is considered a systemic disease with many extra-articular involvements (2), Such as subcutaneous nodules, interstitial lung disease, neuropathy, glomerulonephritis, ischemic heart diseases, etc., primarily observed in longstanding patients (3). RA can also impede physical function and reduce the quality of life drastically in various aspects and aggravate the socio-economic burden on society (4, 5).

Autoimmune thyroid disease (AITD), primarily Graves’ disease and Hashimoto’s thyroiditis, are the leading causes of goiters in non-iodine deficient areas with a prevalence of up to 5% of the population in general. Various autoantibodies are produced in AITD, each targeting specific points in thyroid hormone synthesis (6). AITDs and other systematic autoimmune diseases such as Systemic Lupus Erythematous, Sjögren’s syndrome, Systemic sclerosis, and RA frequently coexist due to apparent pathogenesis (7).

Thyroid dysfunction also seems more prevalent in many autoimmune diseases, including RA (8). Previous studies within different demographics and geographic locations have shown various rates of AITD prevalence among RA patients (9). Although the mechanism of AITDs and RA affecting each other is not yet well defined, several studies have proposed autoimmunity as one of the leading underlying causes contributing to their coexistence and potential influence on each other (6). Considering the potential effect of thyroid abnormality on the severity of RA disease, and few studies accounting racial diversity, it is imperative for further studies to be conducted. Therefore, this study aimed to investigate the association between RA disease severity and thyroid dysfunction, AITD, and anti-thyroid peroxidase (anti-TPO) positivity in the Iranian population.

## Methods

### Study design and participants

In this observational, cross-sectional study, 350 RA patients registered at Shahid Beheshti tertiary care center, Qom, Iran, from April 2021 to March 2022, were included consecutively.

Patients were diagnosed utilizing the American College of Rheumatology and the European League Against Rheumatism (ACR/EULAR) 2010 criteria for rheumatoid arthritis (10). All patients were older than 18, and none suffered from concurrent diabetes mellitus, chronic renal or liver disease, malignancy, current infection, and collagen vascular disease except RA. Patients receiving medication with the potential to cause thyroid test abnormality (e.g., dopamine agonists, interferon-α, anticonvulsant drugs, amiodarone, lithium, and somatostatin analogs). Pregnant women were also excluded.

Before enrollment, all patients gave informed consent, according to the Declaration of Helsinki, and the protocol was approved by the ethics committee residing at medical university of Qom (IR.MUQ.REC.1400.042).

### Clinical assessments

The clinical data were obtained through the patient’s medical record, interview, and physical examination. The data included Patients’ demographic features, prior history of thyroid function disease, duration of RA and thyroid disease, current medications, and the number of tender and swollen joints reported.

Laboratory tests including rheumatoid factor (RF), Erythrocyte sedimentation rate (ESR) using the Westergren method, C-reactive protein (CRP) titer based on Biotec method, mutated citrullinated vimentin autoantibodies (anti-MCV) and anti-cyclic-citrullinated peptide antibodies (anti-CCP) based on an enzyme-linked immunosorbent assay (Elisa) method, Thyroid function tests based on chemiluminescent immunoassay (CLIA) method, and anti-thyroid peroxidase (anti-TPO) antibody using radioimmunoassay were checked. Anti-CCP and anti-MCV were positive for serum levels of more than 5 U/ml and 20 U/ml, respectively. Thyroid function tests included thyroid stimulating hormone (TSH), free thyroxin (FT4), and free triiodothyronine (FT3) with a normal range of 0.3–3.6 mIU/L, 0.7–1.8 ng/dl, 2.57–4.43 pg/mL, respectively. Anti-TPO antibody was considered positive for serum levels of more than 50 IU/mL.

Thyroid dysfunction is defined as abnormal levels of thyroid-stimulating hormone (TSH) with or without abnormal free thyroxin (FT4) or free triiodothyronine levels (FT3) (11). AITD is also described as the simultaneous presence of thyroid dysfunction and anti-TPO antibody. (9)

The disease activity score in 28 Joints for RA with ESR (DAS-28-ESR), which is a tool for measuring the severity of the disease, was calculated with the following formula: (12)

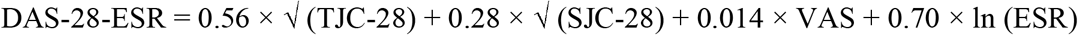

Where TJC and SJC stand for the tender joint count and swollen joint count, respectively, and the visual analog scale (VAS) represents the patient’s global health assessment of disease activity with the score range of 0 as the best to 100 mm as the worst.

DAS-28-ESR values were categorized as follows:

- Remission: DAS-28 ⍰2.6
- mild disease activity: 2.6 < DAS-28 ⍰3.2
- Moderate disease activity: 3.2 < DAS-28 ⍰5.1
- severe disease activity: DAS-28 > 5.1

### Statistical analyses

Quantitate and Qualitative variables were described as mean ± standard deviation (SD) and frequency (percentage), respectively. Kolmogorov-Smirnov test, Q–Q, and P-P plots were used to assess the normality of quantitative variables, in which age and FT3 were normal, and all others were not normal. One-way ANOVA and Kruskal–Wallis test were recruited for normally and non-normally distributed data, respectively. The Chi-square test and ordinal regression were used to compare and analyze categorical variables between different types of disease activity; moreover, the 95% confidence interval (CI) was also reported. Bivariant correlation analysis was used to determine the correlation between quantitative disease activity parameters and thyroid dysfunction test levels. The deletion method was applied where participants had a missing field which was less than 1%. To demonstrate independent factors associated with disease activity, ordinal regression was used. The data analysis was conducted using IBM SPSS Statistics version 26, and the significance of all tests was considered if the P-value < 0.05.

## Results

Two hundred seventy-five females and 75 males with RA were enrolled in this study. By the DAS-28-ESR calculator, 111, 10, 86, and 138 patients were in remission, with mild, moderate, and severe disease activity, respectively. Overall, the patients were sorted into three groups of111 patients in remission, 96 patients with mild-to-moderate, and 138 patients with severe disease activity.

The demographic feature and laboratory findings of all patients are presented in Table 1. Seventy-three out of 112 patients with thyroid dysfunction had a prior history, which all had overt hypothyroidism except for two. ESR, CRP, anti-CCP, and anti-MCV were positive in 11 (10.1%), 14 (12.8%), 60 (54.1%), and 39 (35.5%) patients in remission, 45 (46.9%), 54 (56.3%), 55 (57.3%), and 46 (47.9%) patients with mild-to-moderate disease activity, and 106 (77.4%), 102 (74.5%), 91 (65.9%), and 98 (71.0%) patients with severe disease activity, respectively. As it shown in the table, there was a statistically significant difference between remission, mild-to-moderate, and severe groups for FT3 [F (2, 340) = 35.49, P-value < 0.001] and anti-TPO (H = 15.74, P-value < 0.001).

**Table 1.**
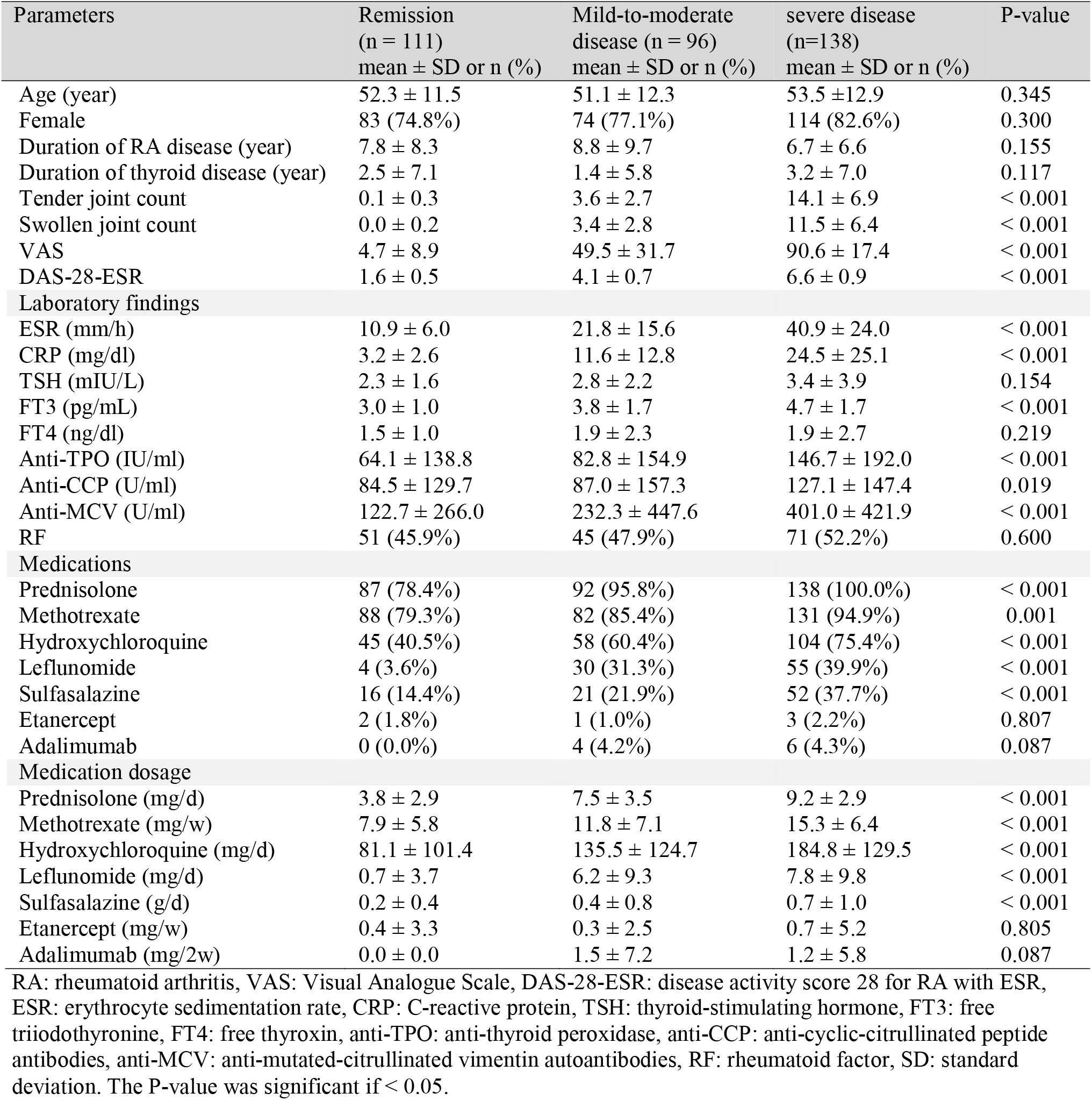
Parameters of inactive and active patients with RA using ANOVA, chi-square, and Kruskal-Wallis test

| Parameters | Remission<br>(n = 111)<br>mean ± SD or n (%) | Mild-to-moderate<br>disease (n = 96)<br>mean ± SD or n (%) | severe disease<br>(n=138)<br>mean ± SD or n (%) | P-value |
| --- | --- | --- | --- | --- |
| Age (year) | 52.3 ± 11.5 | 51.1 ± 12.3 | 53.5 ± 12.9 | 0.345 |
| Female | 83 (74.8%) | 74 (77.1%) | 114 (82.6%) | 0.300 |
| Duration of RA disease (year) | 7.8 ± 8.3 | 8.8 ± 9.7 | 6.7 ± 6.6 | 0.155 |
| Duration of thyroid disease (year) | 2.5 ± 7.1 | 1.4 ± 5.8 | 3.2 ± 7.0 | 0.117 |
| Tender joint count | 0.1 ± 0.3 | 3.6 ± 2.7 | 14.1 ± 6.9 | < 0.001 |
| Swollen joint count | 0.0 ± 0.2 | 3.4 ± 2.8 | 11.5 ± 6.4 | < 0.001 |
| VAS | 4.7 ± 8.9 | 49.5 ± 31.7 | 90.6 ± 17.4 | < 0.001 |
| DAS-28-ESR | 1.6 ± 0.5 | 4.1 ± 0.7 | 6.6 ± 0.9 | < 0.001 |
| **Laboratory findings** |  |  |  |  |
| ESR (mm/h) | 10.9 ± 6.0 | 21.8 ± 15.6 | 40.9 ± 24.0 | < 0.001 |
| CRP (mg/dl) | 3.2 ± 2.6 | 11.6 ± 12.8 | 24.5 ± 25.1 | < 0.001 |
| TSH (mIU/L) | 2.3 ± 1.6 | 2.8 ± 2.2 | 3.4 ± 3.9 | 0.154 |
| FT3 (pg/mL) | 3.0 ± 1.0 | 3.8 ± 1.7 | 4.7 ± 1.7 | < 0.001 |
| FT4 (ng/dl) | 1.5 ± 1.0 | 1.9 ± 2.3 | 1.9 ± 2.7 | 0.219 |
| Anti-TPO (IU/ml) | 64.1 ± 138.8 | 82.8 ± 154.9 | 146.7 ± 192.0 | < 0.001 |
| Anti-CCP (U/ml) | 84.5 ± 129.7 | 87.0 ± 157.3 | 127.1 ± 147.4 | 0.019 |
| Anti-MCV (U/ml) | 122.7 ± 266.0 | 232.3 ± 447.6 | 401.0 ± 421.9 | < 0.001 |
| RF | 51 (45.9%) | 45 (47.9%) | 71 (52.2%) | 0.600 |
| **Medications** |  |  |  |  |
| Prednisolone | 87 (78.4%) | 92 (95.8%) | 138 (100.0%) | < 0.001 |
| Methotrexate | 88 (79.3%) | 82 (85.4%) | 131 (94.9%) | 0.001 |
| Hydroxychloroquine | 45 (40.5%) | 58 (60.4%) | 104 (75.4%) | < 0.001 |
| Leflunomide | 4 (3.6%) | 30 (31.3%) | 55 (39.9%) | < 0.001 |
| Sulfasalazine | 16 (14.4%) | 21 (21.9%) | 52 (37.7%) | < 0.001 |
| Etanercept | 2 (1.8%) | 1 (1.0%) | 3 (2.2%) | 0.807 |
| Adalimumab | 0 (0.0%) | 4 (4.2%) | 6 (4.3%) | 0.087 |
| **Medication dosage** |  |  |  |  |
| Prednisolone (mg/d) | 3.8 ± 2.9 | 7.5 ± 3.5 | 9.2 ± 2.9 | < 0.001 |
| Methotrexate (mg/w) | 7.9 ± 5.8 | 11.8 ± 7.1 | 15.3 ± 6.4 | < 0.001 |
| Hydroxychloroquine (mg/d) | 81.1 ± 101.4 | 135.5 ± 124.7 | 184.8 ± 129.5 | < 0.001 |
| Leflunomide (mg/d) | 0.7 ± 3.7 | 6.2 ± 9.3 | 7.8 ± 9.8 | < 0.001 |
| Sulfasalazine (g/d) | 0.2 ± 0.4 | 0.4 ± 0.8 | 0.7 ± 1.0 | < 0.001 |
| Etanercept (mg/w) | 0.4 ± 3.3 | 0.3 ± 2.5 | 0.7 ± 5.2 | 0.805 |
| Adalimumab (mg/2w) | 0.0 ± 0.0 | 1.5 ± 7.2 | 1.2 ± 5.8 | 0.087 |

RA: rheumatoid arthritis, VAS: Visual Analogue Scale, DAS-28-ESR: disease activity score 28 for RA with ESR, ESR: erythrocyte sedimentation rate, CRP: C-reactive protein, TSH: thyroid-stimulating hormone, FT3: free triiodothyronine, FT4: free thyroxine, anti-TPO: anti-thyroid peroxidase, anti-CCP: anti-cyclic-citrullinated peptide antibodies, anti-MCV: anti-mutated-citrullinated vimentin autoantibodies, RF: rheumatoid factor, SD: standard deviation. The P-value was significant if < 0.05.

Anti-TPO antibody was positive in 18 (16.2%) patients in remission, 25 (26.9%) patients with mild-to-moderate disease activity, and 58 (43.6%) patients with severe disease activity. Among patients with severe disease activity, it was 2.93 times more probable to detect anti-TPO positive value than the patients in remission (OR =2.93, 95% CI: 1.86 - 4.62; P-value < 0.001). The number of patients with thyroid dysfunction and AITD were 27 (24.3%) and 13 (11.7%) in remission, 27 (28.1%), and 10 (10.4%) mild-to-moderate disease activity, and 55 (40.1%) and 37 (28.0%) with severe disease activity, respectively. Hence, the patients with higher disease activity were 82% more likely to have thyroid dysfunction (OR = 1.82, 95% CI: 1.19 - 2.77; P-value = 0.006) and 2.7 times more probable to have AITD than the remission patients (OR = 2.71, 95% CI = 1.57 – 4.62; P-value < 0.001).

DAS-28-ESR level was positively correlated with anti-TPO (r = 0.192; P-value < 0.001) and FT3 (r = 0.388; P-value < 0.001), but there was no significant correlation with TSH nor FT4. The dose of Methotrexate and Hydroxychloroquine was only correlated with FT3 (r = 0.371; p-value < 0.001 and r = 0.121; p-value = 0.024, respectively). Yet, Prednisolone dosage was positively correlated with FT3 (r = 0.404; p-value < 0.001) and anti-TPO (r = 0.236; p-value < 0.001) levels but did not have any significant correlation with FT4 and anti-TPO level. Predictive factors of disease severity are shown in Table 2. As it showed higher levels of thyroid function tests, anti-TPO, and female gender were significantly associated with higher RA disease activity.

**Table 2.** Ordinal regression analysis determining independent predictors affecting rheumatoid arthritis disease severity

| Variables | OR | P-value |
| --- | --- | --- |
| Age | 1.008 | 0.370 |
| Gender (female sex) | 1.820 | 0.025 |
| Duration of thyroid disease (year) | 0.977 | 0.198 |
| TSH (mIU/L) | 1.117 | 0.027 |
| FT3 (pg/mL) | 1.696 | < 0.001 |
| FT4 (ng/dl) | 1.153 | 0.011 |
| Anti-TPO (IU/ml) | 1.002 | 0.046 |

TSH: thyroid-stimulating hormone, FT3: free triiodothyronine, FT4: free thyroxine, anti-TPO: anti-thyroid peroxidase. The P-value < 0.05 was significant if < 0.05.

## Discussion

The coexistence of RA with AITD has been long debated due to similar pathogenesis. However, a consensus on this matter is not achieved (13). Several studies have established a higher prevalence of thyroid dysfunction, particularly overt hypothyroidism, anti-TPO positivity, and AITD among RA patients (14, 15). In addition, recent studies have also suggested that AITD comorbidity can exacerbate RA severity and activity (16). On the contrary, Joshi et al. found high RA disease activity was more common among patients with hypothyroidism, though it was not statistically significant (17). Also, some studies have found no association between thyroid disease and RA disease activity which is in contrast with the findings of the aforementioned studies (18).

Due to conflicting results on this matter, we aimed to assess the association between RA severity and thyroid dysfunction, anti-TPO positivity, and AITD. In our study, 350 RA patients were enrolled and were divided into three sub-groups based on disease severity which was determined based on the DAS-28-ESR score. The first group consisted of 111 patients in remission, the second group included 96 patients with mild-to-moderate disease activity, and the last group contained 138 patients with severe disease activity. In each group, the clinical and paraclinical factors determining the severity of RA were analyzed regarding serum levels of thyroid markers and thyroid auto-antibodies. Our study’s data analysis yielded a significant association between RA disease severity and thyroid dysfunction, AITD, and anti-TPO positivity. Higher RA disease severity was also significantly associated with higher serum TSH, FT3, FT4, and anti-TPO levels and the female gender.

Recent studies have shown that RA patients with hypothyroidism or positive anti-TPO had higher DAS-28-ESR levels (19).In addition, Koszarny et al. found a positive correlation between DAS-28-ESR and anti-thyroid antibodies, including anti-TPO levels (20). However, Atzeni et al. found no correlation between anti-thyroid antibodies and RA disease activity parameters (21). In our study, DAS-28-ESR was positively correlated with FT3 and anti-TPO serum levels. Elattar et al. demonstrated that TSH level was positively correlated with RA disease activity parameters, including Methotrexate dosage (16). Although in our study, no correlation was found between the dose of Methotrexate and TSH serum level, a remarkable positive correlation between Methotrexate moreover Hydroxychloroquine dosage and FT3 serum level were found. Consumption dosage of other medications such as Prednisolone was also positively correlated with FT3 and anti-TPO serum levels, but no significant correlation with other thyroid function tests was found.

Noteworthy, our study revealed that the patient’s age had no significant impact on disease activity which may have been due to the insufficient sample size. Another shortcoming of this study that leaves room for further studies on this matter was not measuring other anti-thyroid autoantibodies and their correlation with RA activity.

In conclusion of our study, thyroid abnormality, including thyroid dysfunction and AITD comorbidity with RA disease, had a considerable impact on the RA disease severity, which can affect the quality of life and prognosis of the disease and widens the scope of approaching RA management and its numerous complications.

## Data Availability

The datasets generated and analyzed during the current study are not publicly available due to restrictions in ethical and data management approvals. Still, they are available from the corresponding authors upon reasonable request.

## Conflicts of interest

The authors declare that there is no conflict of interest regarding the publication of this article.

## Ethical Approval

The study was conducted according to the Declaration of Helsinki and the protocol approved by the ethics committee of Qom University of Medical Sciences (IR.MUQ.REC.1400.042).

## Funding statement

Not applicable.

## Authors’ contributions

MM and MB designed the study. MBF and MAY wrote the proposal, and all authors contributed to preparing the final draft. MM and MAY collected the data. MV and MBF analyzed the data. MAY, MBF, and MAH wrote the manuscript, and all authors critically revised the manuscript and approved the final manuscript.

## Acknowledgments

Not applicable.

## Notes

### Competing Interest Statement

The authors have declared no competing interest.

### Funding Statement

This study did not receive any funding

